# Home-based High Intensity Functional Strength Training (HIFST) for Older Adults: A Protocol for a Pilot Randomized Controlled Trial and Qualitative Description of an Exercise Program to Prevent Functional Decline after an Injury

**DOI:** 10.1101/2022.09.13.22279895

**Authors:** Ashley Morgan, Ada Tang, Jennifer Heisz, Lehana Thabane, Julie Richardson

**Affiliations:** School of Rehabilitation Sciences, McMaster University, Institute of Applied Health Sciences, 1400 Main Street West, Hamilton, Ontario, Canada, L8S 1C7; Department of Kinesiology, McMaster University, Ivor Wynn Centre, 206, 1280 Main Street West, Hamilton, Ontario, Canada, L8S 4K1; Department of Health Research Methods, Evidence, and Impact (HEI), McMaster University Medical Centre, 1280 Main West 2C Area, Hamilton, Ontario, Canada, L8S 4K1,; St Joseph’s Healthcare, Hamilton, Hamilton ON, Canada; Faculty of Health Sciences, University of Johannesburg, Johannesburg, South Africa

**Keywords:** pilot study, mobility limitation, high-intensity exercise, older adults, tele-rehabilitation

## Abstract

**Background:** Slip, trips, and falls are a common cause of injury in older adults; limiting physical activity participation and mobility task performance. These injuries may result in preclinical mobility limitation (PCML), a period in which individuals report modifications but not difficulty in mobility tasks. Individuals with PCML are at risk of functional decline which can be prevented with exercise. High-intensity functional strength training (HIFST) involves short intervals of ‘hard’ interspersed with ‘easy’ exercise that may be a time efficient strategy to improve functioning for older adults experiencing PCML.

**Objective:** This protocol outlines the rationale, methods, and planned analyses for a pilot randomized controlled trial and qualitative description (QD) to investigate the feasibility, preliminary effects, and acceptability of a home-based 12-week HIFST intervention for community-dwelling older adults (≥ 55 years) with PCML who have had an injury from a slip, trip, or fall.

**Methods:** Twenty-four participants (target) will be randomized into a 12-week home-based HIFST or lower extremity stretching intervention. Feasibility will be determined using criteria for adherence, recruitment, retention, and safety and results will be presented using descriptive statistics. Preliminary effects on physical functioning, cognitive functioning, enjoyment, and harms will be assessed and presented as mean between-group differences with 95% confidence intervals. HIFST participants will be recruited for follow-up interviews using QD methodology to investigate the acceptability of the intervention. The results of this pilot trial will provide essential information for future research regarding the process, resources, and scientific merit of conducting home-based high-intensity exercise in a post-injury older adult population.

**Trial Registration:** NCT05266911

## Introduction

Previous estimates suggest that approximately 15% of Canadians have sustained an activity-limiting injury in the past year.^1^ Falls are the most common cause of injury hospitalization in Canada and more than half of activity-limiting injuries in older Canadians are the result of a fall, most often from tripping or stumbling while walking or doing household chores.^1^ An injury resulting from a slip, trip or fall in an older adult may decrease self-efficacy,^2^ restrict mobility,^2,3^ and increase the risk of functional decline.^2,3^ An individual’s level of functioning depends on various factors, including adequate physical and cognitive functioning,^4^ and exists on a continuum which can include periods of decline and restoration of function.^4,5^ Preclinical mobility limitation (PCML) is a period located along this continuum characterized by subtle modifications in frequency or method of task performance that are indicative of an early stage of functional decline.^6,7^ An injury from a slip, trip, or fall in an older adult may initiate the onset of PCML. However, while many falls-specific programs exist,^8^ this post-injury period has received minimal attention for exercise interventions.^9^

High intensity functional strength training (HIFST) can be defined as a “training style [or program] that incorporates a variety of functional movements, performed at high-intensity [relative to an individual’s ability.]”^10(p2)^ High intensity interval training (HIIT), which involves brief intense bouts of aerobic activity combined with rest or low intensity exercise has been shown to be feasible, safe, and beneficial for cardiovascular function in older adults.^11^ Many HIFST programs apply the HIIT interval format to functional strengthening exercises (usually multi-joint movements like sit-to-stands) in intense work to rest/recovery ratios.^10,12^ By utilizing an interval format during HIFST, heart rate is elevated and there is potential to gain benefits from both aerobic and resistance training.^10,12^

Despite, these benefits, HIFST has not received as much attention in the literature as HIIT and few studies have examined it’s impact on both physical and cognitive functioning.^13^ Physical and cognitive functioning impairments often co-exist and increase the risk for falls and functional decline.^14^ Specifically, deficits in the cognitive domains of executive functioning (complex cognitive abilities related to the planning and execution of goal-directed behaviours, abstract reasoning, and judgement^15,16^)^15,16^ and processing speed, in particular have been linked with poorer mobility outcomes,^14^ including higher risk of falls and reductions in walking speed.^17^

Additionally, evidence suggests that enjoyment is an important, but understudied factor which may contribute to adherence^18,19^ and acceptability of physical activity interventions for older adults.^20^ Acceptability is a multi-faceted construct, reflecting the extent to which people delivering or receiving an intervention consider it to be appropriate.^21^ The acceptability of an intervention to a participants can be influenced by the content, context, and delivery.^21^ The theoretical framework of acceptability of healthcare interventions outlined by Sekhon et al., considers the following component constructs; affective attitude (feelings toward intervention), burden (perceived effort to participate), ethicality (fit with value system), intervention coherence (understanding of intervention and how it works), opportunity costs (benefits, profits or values given up), perceived effectiveness (perception that the intervention will achieve its purpose) and self-efficacy (confidence in performing required behaviours).^21^ Acceptability can be explored using quantitative and qualitative methods,^21^ which both add unique and valuable information. The inclusion of qualitative data in pilot trials can be instrumental in determining and correcting potential problems that can impact the acceptability and implementation of an intervention.^22^

To our knowledge, no previous research exists examining the feasibility (e.g., adherence, recruitment, retention, safety), effects (physical functioning, cognitive functioning, or enjoyment) or acceptability of a home-based HIFST program for community-dwelling older adults who are post-injury from a slip, trip, or fall. Thus, we propose a pilot study to measure the feasibility of a home-based HIFST program, its preliminary effects on physical and cognitive functioning, changes in physical activity enjoyment and the acceptability of the intervention for participants. This proposed pilot trial will combine collection of quantitative data in a randomized controlled trial (RCT) with a follow-up qualitative description (QD) study.

### Purpose

The purpose of the pilot study is to assess the feasibility, preliminary evidence of effects, and acceptability of a home based HIFST program delivered by a physiotherapist (PT) to community-dwelling older adults (≥ 55 years) after an injury from a slip, trip, or fall.

The primary and secondary research questions for the pilot RCT are:

1. Primary: Is 12-weeks of home-based HIFST for community-dwelling older adults (≥ 55 years) post-injury feasible as determined by adherence, recruitment, retention, and safety criteria? **Criteria:**
  1a) Adherence: ≥ 70% of total sessions completed^13,23–25^
  1b) Recruitment: To recruit ≥ 65% of eligible participants^24,26,27^ and outline details regarding recruitment source, and reasons for exclusion.
  1c) Retention: ≥ 80 % of participants enrolled will complete intervention.^13,23,25^
  1d) Safety: No occurrence of any intervention-related major adverse event.
2. Secondary: Do community-dwelling older adults who are post-injury and receive 12 weeks of HIFST show preliminary evidence of effects on measures of physical functioning, cognitive functioning, and physical activity enjoyment?
  2a) Physical functioning: 4 meter (m) walk test,^28^ 2 minute step test,^29^ dual-task cognitive timed up and go (TUG-COG),^30,31^ 30-second chair stand (30CST)^29^ and a Mänty PCML scale.^32^
  2b) Cognitive functioning: Executive functions and processing speed via California Older Adult Stroop Test (COAST),^33^ Oral Trail Making Test,^34,35^ and Digit Symbol Substitution Test (DSST).^36,37^
  2c) Enjoyment: Physical Activity Enjoyment Scale (PACES)^38^ and Feeling Scale(FS).^39^
  2d) Harms: any intervention-related adverse events (e.g., muscle soreness, muscle strains, joint pain) reported by participants.

The primary research question for the qualitative description is:

What is the acceptability to participants of home-based HIFST delivered by a physiotherapist to community-dwelling older adults (≥ 55 years) who are at risk for functional decline following an injury?

## Methods

This protocol follows the Standard Protocol Items: Recommendations for Interventional Trials (SPIRIT) checklist^40^ and is informed by the Consolidated Standards of Reporting Trials (CONSORT) statement extension to randomised pilot and feasibility trials^41^ and recommendations for pilot trials presented by Thabane et al.,^42^ This research did not receive any specific grant from funding agencies in the public, commercial, or not-for-profit sectors.

### Study Design

The pilot parallel-group RCT will be single blinded (outcome analysis) as the nature of the exercise intervention prevents participant and therapist blinding. The qualitative study will involve semi-structured interviews post-intervention with participants randomized to the HIFST program. Qualitative description methodology, which aims to create a rich, straight description of an experience or event,^43^ will be utilized. It presents the results in everyday language similar to that utilized by the participants^43,44^ and is well suited to providing practice answers to the who, what and where of an experience or event.^44^ This trial is registered on clinicaltrials.gov; NCT05266911. A study timeline can be found in Figure 1.

**Figure 1.**
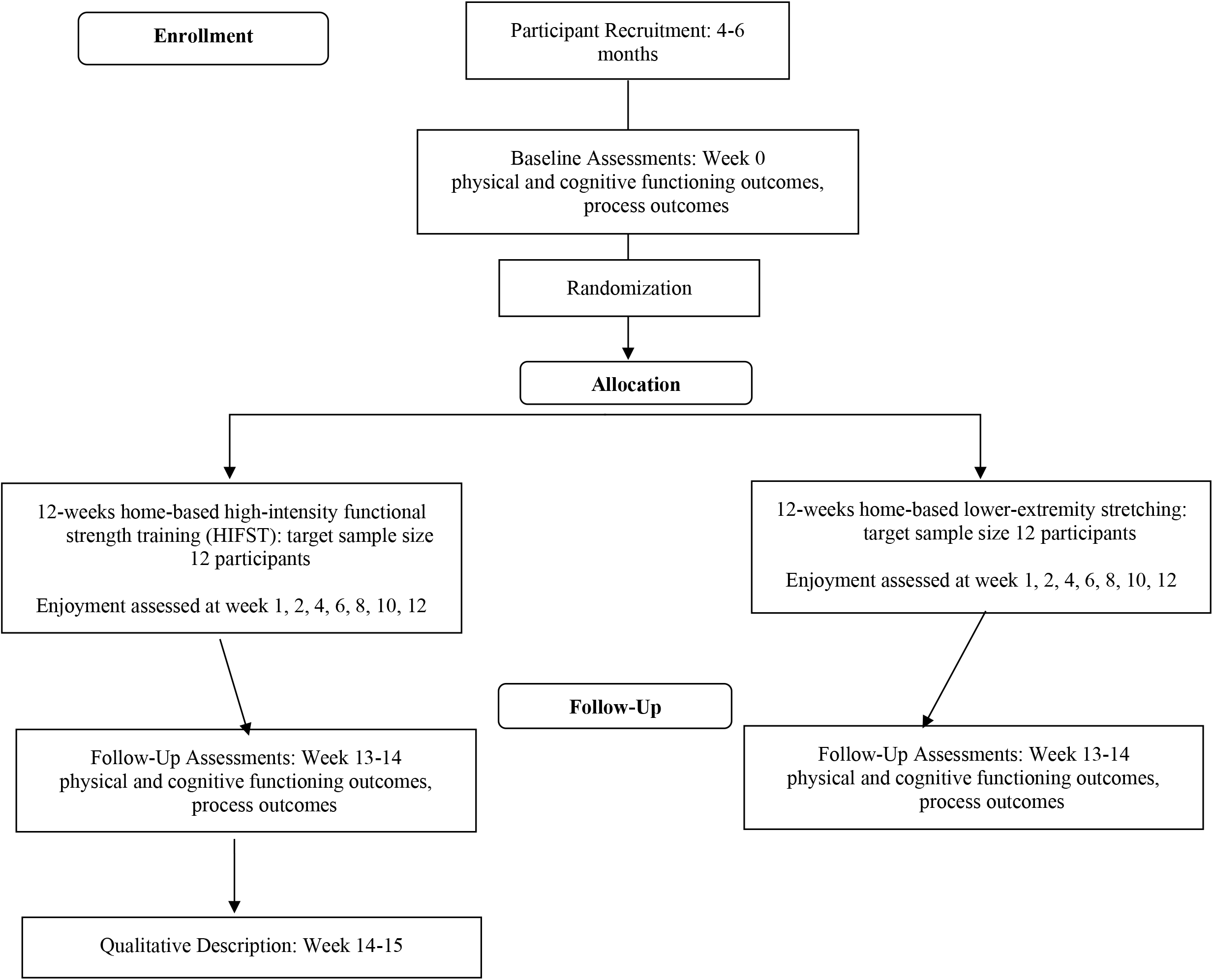
Study Timeline.

### Participants

For the RCT, Participants will be screened for participation via telephone, based on the following inclusion and exclusion criteria.

#### Inclusion criteria

Eligible participants will include 1) English-speaking, community-dwelling older adults ≥ 55 years, who 2) sustained an injury from a slip, trip or fall in the last six months (assessed by self-report) and 3) report decreased and/or modified daily task performance (assessed using PCML questionnaire based on Mänty.)^32^ Participants must have 4) no contraindications to exercise based on the American College for Sports Medicine recommendations^46,47^ and 5) complete the Canadian Society for Exercise Physiology ‘Get Active’ questionnaire^48^ and obtain clearance from a health care professional if deemed necessary based on screening.^48^ 6) Participants will be required to have access to email and a laptop/tablet with webcam capable of running the web-based videoconferencing platform Zoom well as 7) the ability to provide informed consent.

#### Exclusion criteria

Participants will be excluded if they have a score of < 11 on the Mini Montreal Cognitive Assessment (Mini MoCa).^49^

For the follow-up QD, participants randomized to the HIFST intervention will be eligible and invited to participate in the interviews.

### Recruitment

Participants will be recruited from Hamilton, Ontario, Canada, and surrounding communities in southern Ontario. Participants will be recruited from the community utilizing advertisements in community physical or virtual spaces, print media and through various community organizations/groups. If recruitment is slow after 1-2 months we will consider additional strategies, such as social media advertising and recruiting through healthcare settings such as rehabilitation or medical clinics. We will attempt to recruit equal numbers of men and women. A rolling recruitment strategy be utilized on an ongoing basis until the required sample size (24) is reached (see Figure 1).

### Sample Size

Based on published recommendations for pilot studies^42,50,51^ sample size for the RCT was established using a confidence interval (CI) based on the primary outcome of feasibility as measured by adherence rates (see calculation Appendix A). Accounting for a 20% drop out, the target sample size is 24 participants (12 in each group).

### Randomization

Eligible participants who provide informed consent will be stratified by sex and allocated to the intervention (HIFST) or control (lower extremity stretching) group on a 1:1 ratio using variable block sizes of 2, 4, and 6, after the completion of the baseline assessment. The random allocation sequence will be computer generated through the online program Sealed Envelope Ltd.,^52^ and uploaded to a web-based randomization system (Research Electronic Data Capture [REDCap])^53^ by an individual uninvolved in trial administration. REDCap^53^ will be utilized for group allocation which will be concealed until the time of randomization.

### Proposed methods for protecting against sources of bias

#### Blinding

Participants will be blinded to their group allocation at baseline assessment. During the intervention, it will not be possible to blind the participants or physiotherapist (PT) delivering the intervention. Outcome assessment will not be blinded at follow-up assessments. Outcome analysis will be blinded to group allocation.

#### Contamination

The intervention and control group will be delivered via internet videoconferencing (Zoom)^54^ at each participant’s home by a PT which limits the opportunity for contamination. Each participant will be provided with a link to the secure videoconferencing platform which will only be available to them. Participants in both groups will be asked to refrain from beginning a new exercise program. If participants are receiving active rehabilitation treatment, the details (frequency, components) will be documented.

### Trial interventions

Participants will be randomized to either a home-based high intensity functional strength training group (intervention) or a lower extremity stretching (control) group. The details of both interventions, utilizing the Consensus for Exercise Reporting Template (CERT)^55^ criteria are found in Table 1.

**Table 1.** Intervention Descriptions using CERT criteria.

| **Category** | **Experimental intervention:**<br>**HIFST program** | **Control: Lower extremity**<br>**stretching** |
| --- | --- | --- |
| **BRIEF DESCRIPTION** | Home-based program of intervals of 'hard' effort (Borg's rating of perceived exertion [RPE]) of strengthening exercises interspersed with 'easy' activity. | home-based program of lower extremity stretching |
| **WHAT:** materials (exercise equipment) | Focus on bodyweight exercises with minimal/no equipment. Household objects (e.g., chair, laundry basket) or available exercise equipment (e.g., dumbbells) will be employed as/if needed. | May need chair and towel/rope to assist with stretches. |
| **WHO:** provider (exercise delivery) | Using videoconferencing, a physiotherapist (PT) will provide initial exercise prescription and supervision, 2 additional individually supervised sessions (week 1-2) and 2 supervised virtual group sessions (week 3-4). Week 6, 8, 10, 12, the PT will have virtual meetings with participants for progression/modification. | Using videoconferencing, a PT will provide initial session of stretching exercise prescription and supervision. Participants will have a virtual meeting with the PT at week 4 and 8. |
| **HOW:** delivery (group/individual) | All individual except for 2 virtual small group ($\leq 4$ participants) sessions which are for monitoring purposes only. | Individual |
| **HOW:** delivery (supervised/unsupervised) | 3 initial individual virtual sessions and 2 virtual group sessions supervised; during which time participants are expected to also be completing program independently on an additional 1-2 days/week. Remaining sessions to be completed unsupervised with biweekly virtual meetings with PT. | 1 initial supervised session and remaining unsupervised with virtual meetings with PT at week 4 and 8. |
| **HOW:** delivery (measurement and adherence reporting) | Attendance will be recorded at supervised sessions and participants will keep an exercise log. Logs will be emailed to PT prior to each virtual meeting. | Attendance will be recorded at supervised session and participants will keep an exercise log. Logs will be emailed to PT prior to each virtual meeting. |
| **HOW:** delivery (motivation strategies) | Graduated approach to intensity and increased independence. Use of periodic meetings, email reminders, and exercise logs. | Periodic meetings, email reminders, and exercise logs. |
| **HOW:** delivery (decision rules for progression) | Intensity will be progressed gradually over the first 4 weeks and length of interval time and numbers of rounds will also be progressed (e.g., week 1: 2 rounds of 20 seconds 11-13 RPE, 1 minute $\leq 9$; week 12: 4 rounds of 30 seconds 15-17 RPE, 30 seconds $\leq 9$) Exercise progression based on ability to achieve target RPE (15-17) with proper execution. Progressions made at supervised sessions or virtual meetings. | Lower extremity stretches can be progressed (e.g., increase range-of-motion, change position) at virtual meetings if needed. |
| **HOW:** delivery (exercise description) | *Warm-up:* walking in place, side stepping, marching, marching with overhead reach, etc.,<br>*Work intervals:* Lower extremity focused functional compound movements based on 1. sit to stand, 2. stair stepping, 3. lunging, 4. lifting/carrying<br>*Recovery intervals:* slow walking, side stepping, marching<br>*Cool-down:* 3-5 minutes easy activity (walking in place, marching) followed by stretching | *Lower extremity stretches:* standing lunge stretch, standing calf stretches, seated/supine hamstrings, standing or lying quadriceps, kneeling, or standing thoracic/lumbar (back) stretch |
| **HOW:** delivery (home program content) | 100% home-based | 100% home-based |
| **HOW:** delivery (nonexercise components) | education: using RPE scale, safety considerations and planning, exercise logs and adherence, last session: long-term exercise plans | education: safety considerations and planning, exercise logs and adherence (verbal and written) |
|  | and suggestions for future modifications/progressions (verbal and written) |  |
| **HOW:** delivery (adverse event documentation and management) | Participants will be informed of all potential safety risks and will also receive education regarding indications to discontinue/modify exercise, and expectations and management for post-exercise soreness. Participants will be emailed weekly reminders of exercise monitoring, self-screening, and documentation. All adverse events will be documented in a standardized form and obtained via participant report (exercise log, email contact, virtual meetings) or PT report (supervised sessions). We will ensure that participants understand the steps to take if a suspected adverse event should occur and will be required to identify an emergency contact. We will also provide them with an email contact to report an adverse event and ask about any events at each meeting with the PT. | Participants will be informed of all potential safety risks and will also receive education regarding indications to discontinue/modify exercise, and expectations and management for post-exercise soreness. Participants will be emailed weekly reminders of exercise monitoring, self-screening, and documentation. All adverse events will be documented in a standardized form and obtained via participant report (exercise log, email contact, virtual meetings) or PT report (supervised sessions). We will ensure that participants understand the steps to take if a suspected adverse event should occur and will be required to identify an emergency contact. We will also provide them with an email contact to report an adverse event and ask about any events at each meeting with the PT. |
| **WHERE:** location | participants' homes | participants' homes |
| **WHEN, HOW MUCH:** dosage | 3 days/week; approximately 30-35 minutes<br>4 exercises completed in an interval format with number of rounds, intensity and interval duration progressed over time. Target "high"/hard intensity of 15-17 RPE reached by week 4. | 3 days/week; approximately 15 minutes<br>5 stretches; 30 seconds per side, 3 rounds |
| **TAILORING:** (what, how) | Specific exercises will be tailored to individual's ability by modifying range-of-motion, speed, and/or external resistance | Specific stretches will be tailored to individual's ability (modify range-of-motion, position) |
| **HOW WELL:** planned, actual | Participants will report completion of exercises and exercise parameters (rounds, time, intensity achieved) in exercise logs. | Participants will report completion of stretches and duration in exercise logs. |

### Safety Considerations

The intervention will be tailored to each participant’s ability to minimize the risk for adverse events. Information related to patient safety education and adverse event documentation can be found in Table 1. Systematic reviews examining high-intensity exercise in older adults suggest that the risk of adverse events is low.^11,13,56^ The most common adverse events that individuals who participate in exercise interventions may experience are musculoskeletal related, such as muscle soreness, muscle strains, joint pain or falls.^57^ To minimize risks, all participants will be screened for contraindications to participation^46,47^ to prevent enrollment of individuals who do not meet the inclusion criteria. Eligible participants will meet with the PT for instruction and supervision of a tailored intervention which will be monitored and progressed gradually by the PT.

### Outcomes: RCT

#### Primary Outcomes

As a pilot trial, the primary outcome is **feasibility** determined by **adherence, recruitment, retention, and safety criteria**. 1. **Adherence** is the primary outcome measure for feasibility. An acceptable adherence rate is set at ≥ 70% of all sessions completed based on literature reporting adherence rates for exercise,^25,56^ and higher intensity exercise in older adult populations.^13,23^ Attendance will be recorded by the PT for the supervised sessions and self-reported by participants in an exercise log and emailed to PT prior to each virtual meeting. 2. **Recruitment;** The percentage of eligible participants recruited will be ≥ 65% to meet feasibility criteria based on similar interventions^24,58^ and target populations.^26,27^ Recruitment criteria will be assessed by keeping records logging date of recruitment, source, and reasons for exclusion. 3. **Retention** aims to be ≥ 80% based on the available literature^13,23,25^ and will be assessed by calculating the percentage of participants who complete both baseline and follow-up assessments. 4. **Safety;** To meet safety criteria the trial must demonstrate no serious adverse events (e.g., life threatening, require hospitalization or cause significant disability)^59^ related to the exercise intervention. No serious adverse events are expected based on the results from similar interventions.^11,13,23,24,58^ Safety criteria will be assessed via participant report (exercise log, email contact, or at virtual PT meetings) which will be documented in a standardized safety form.

#### Secondary Outcomes

Secondary outcomes will be preliminary measures of effects on **physical functioning, cognitive functioning, and physical activity enjoyment** as well as any **harms** reported. **Harms** will be recorded throughout the intervention and assessed via participant report as described previously. Any serious adverse events will be considered under the feasibility criteria and all other minor events (e.g., muscle soreness, etc.,) will be reported under the secondary outcome of ‘harms’. A full description of each outcome measure, including relevant measurement properties can be found in Appendix B. Physical functioning and cognitive functioning outcomes will be assessed prior to randomization and after the 12-week intervention (see study timeline) using a secure videoconferencing platform (Zoom).^54^ Physical activity enjoyment will be assessed via self-report (email) throughout the 12-week intervention (see study timeline).

**Physical functioning** will be measured utilizing the 4 meter (m) walk test (assess usual gait speed),^59^ 2 minute step test (TMST, assess exercise capacity)^29^ dual-task cognitive Timed Up and Go (TUG-COG, assess dual-task mobility),^30,31^ 30-second chair stand test (30CST: assess lower body strength),^29^ and a Mänty PCML scale (assess PCML status).^32^ **Cognitive functioning**, specifically processing speed and the executive functions of inhibitory control, task-shifting (mental flexibility), and attention will be assessed utilizing the California Older Adult Stroop test (COAST, assess inhibitory control),^33^ Oral Trail Making Test (OTMT, assess task-shifting, attention, working memory and processing speed),^34,35^ and Digit Symbol Substitution Test (DSST, assess processing speed and several aspects of executive functioning).^36,37^ Evidence suggests these aspects of cognition are associated with mobility outcomes, including gait performance.^14^ **Physical activity enjoyment** and affective response to exercise will be measured by the 8-item Physical Activity Enjoyment Scale (PACES)^38^ and the single-item Feeling Scale (FS)^39^ at 7 timepoints throughout the intervention (see study timeline).

#### Process Outcomes

Self-efficacy for exercise, using the Self Efficacy for Exercise Scale (SEE)^62^ and the Activities Balance Confidence (ABC) Scale^63^ will also be measured at baseline and post-intervention.

### Outcomes: Qualitative Description

Data generation will occur following the 12-week HIFST intervention period using semi-structured in-depth interviews via videoconferencing (Zoom)^54^ and an online anonymous survey with open-ended questions. To contextualize the findings, a brief questionnaire regarding participant demographics, weeks of intervention participation, adherence, and technical difficulties encountered will be collected via questionnaire prior to the interviews. A draft interview guide and online survey can be found in Table 2. The interview guide development was influenced by the framework of acceptability proposed by Sekhon et al.,^21^ The interview guide will provide an overarching structure for the interviews but is somewhat flexible and the use of additional probes or order of questions may vary with each participant. Field notes will be taken during and following interviews by AM and will be utilizing as an additional data source. These notes will include observations, areas to probe further and highlight key details or ideas.

**Table 2.** QD Draft Interview Guide and Questionnaire.

| Question | Prompts | Related Constructs from Acceptability of Healthcare Interventions framework<sup>21</sup> |
| --- | --- | --- |
| 1. For 12 weeks you were involved in a high intensity functional strength training program. Can you describe your overall experience to me? |  |  |
| 2. Can you please explain how you heard about the study and your reasons for joining? | - What were your expectations prior to beginning the program?<br>- How did the program meet or not meet these expectations?<br>- What did your friends/family think about your participation? | affective attitude, self-efficacy, intervention coherence |
| 3. Prior to participation in this study, can you describe your physical activity level? | - Would you say you have experienced changes to activity in the last 5 years, and why do you think this is?<br>- What did you think or know about high-intensity functional strength exercise prior to participating in this study?<br>- Can you describe any experiences participating in high intensity or vigorous activity? | affective attitude, self-efficacy, intervention coherence |
| 4. What did you think about the overall delivery of the program? | - How did you find the virtual aspects of the program?<br>- What did you think about the effectiveness of the PT instructor?<br>- Was there anything else you specifically liked or disliked? | affective attitude, perceived effectiveness, |
| 5. How did you find the content of the program? | - What did you think about the specific exercises? High-intensity interval format?<br>- How did you feel completing the exercises and did this change over time? (affect) | affective attitude, perceived effectiveness |
| 6. Can you please describe any factors that made completing the program more challenging for you? (barriers) (OR if discontinued: Can you please describe your reasons for discontinuing the program?) | - Can you please describe any strategies utilized to overcome challenges?<br>- Would you say it required a good deal of effort or very little effort to complete the program as prescribed? Why do think so? | burden, opportunity costs |
| 7. Can you please describe any factors that encouraged or made participation in the program easier?(facilitators) | - Did any of these factors change over time, and if yes, how so? | self-efficacy |
| 8. Can you describe whether you feel like you experienced any changes in your physical or cognitive function from participating in the program? | - Do you notice any difference in how you complete your daily activities compared to before?<br>- Did you experience any negative effects? | perceived effectiveness, burden, opportunity costs |
| 9. Now this trial is completed, can you describe any plans you have for future physical activity? | - What aspects of this type of activity are appealing?<br>- Do you think you will continue to participate in high-intensity exercise? Why or why not? | self-efficacy, intervention coherence, perceived effectiveness, affective attitude |
| 10. The next step is to evaluate this intervention in a larger study. What components would you recommended need to be adapted or changed? Stay the same? |  |  |
| **Online Questionnaire** |  |  |
| 1. What did you think about the overall delivery of the program (including instructor)?<br>2. What did you think about the content of the program?<br>3. Please describe any factors which influenced your participation in the program and whether these increased or decreased participation.<br>4. Please explain why you plan (or do not plan) to continue to participate in high-intensity exercise (or exercise in general).<br>5. Please provide any additional feedback, including components of the intervention you think should be changed. |  | affective attitude, burden, intervention coherence, opportunity costs, perceived effectiveness, self-efficacy |

**Table 3.** Pilot RCT Analyses Summary.

| Variable | Outcome Measures | Criteria for success/Hypothesis | Method of Analysis |
| --- | --- | --- | --- |
| **Primary** |  |  | Descriptive statistics |
| Recruitment | Percentage (%) of eligible participants recruited | ≥ 65% of eligible participants recruited |  |
| Retention | % of participants completing both baseline and follow up assessments | ≥ 80% of participants enrolled |  |
| Adherence* | % of all sessions completed | ≥ 70% of all sessions completed |  |
| Safety | Record of serious intervention-related adverse events | No serious adverse events related to exercise intervention |  |
| **Secondary** |  |  |  |
| Physical Functioning | - 4-meter walk test<br>- 2-minute step test<br>- dual-task cognitive Timed Up and Go<br>- 30-second chair stand test<br>- Mänty preclinical mobility limitation scale | Improvements in intervention group relative to control group | t-tests or Wilcoxon-Mann-Whitney tests |
| Cognitive Functioning | - California Older Adult Stroop test<br>- Digit Symbol Substitution Test<br>- Oral Trails Making Test | Improvements in intervention group relative to control group | t-tests or Wilcoxon-Mann-Whitney tests |
| Enjoyment | - Physical Activity Enjoyment Scale (PACES)<br>- Feeling Scale (FS) | Assess trajectory of change in enjoyment in both groups and differences between groups (greater enjoyment in intervention group). | mixed effects modelling |
| Harms | Record of all intervention-related adverse events | No difference between groups | Descriptive statistics |

\* Adherence is the primary feasibility outcome

### Outcomes: Criteria for Progression to a Future Definitive Trial

To proceed to a future RCT, the four feasibility criteria outlined should be met, preliminary evidence of positive effects in at least one physical or cognitive outcome measure demonstrated, and the results of the qualitative study indicative of acceptability among the participants. If any of these criteria are not met, considerations about modifications that could be made to the protocol to positively impact unmet criteria will be made. The recommendations outlined by Bugge et al., for decision-making after pilot and feasibility trials will be employed to guide this process.^64^

### Quantitative Analysis

We will adopt the CONSORT extension to pilot trials^41^ in reporting the results of the pilot trial. A summary of analyses can be found in Table 2. All statistical calculations will be made utilizing the software Stata IC/16.0 (StataCorp LLC, Texas USA). Participant baseline characteristics (see Appendix II for demographic questionnaire) will be analyzed using descriptive statistics and reported for continuous variable as means and standard deviations given a reasonably normal distribution and as medians and quartiles for skewed distributions. Counts and percentages will be used to describe categorical data.

The primary outcomes assessment of **feasibility** will be analyzed using descriptive statistics. Adherence will be reported as the percentage of total intervention (HIFST) sessions completed. Recruitment rates will be determined by the percentage of eligible participants who are subsequently enrolled in the study. Retention rates will be reported as the percentage of participants enrolled who complete the intervention. Adverse events will be reported as counts of minor or serious events with a description of the event and the group allocation. Data will also be presented disaggregated by sex and gender to explore any differences in the feasibility criteria.

This pilot RCT does not aim to, nor is powered to, assess effectiveness of the HIFST program. However, to inform a future trial, exploratory statistical analysis will be conducted to assess the preliminary effects on secondary outcome measures. For all physical and cognitive functioning measures (see Appendix I) between group differences will be analyzed using t-tests if assumptions of normality and homogeneity of variance are met or the Wilcoxon-Mann-Whitney test if they are not met. Results will be expressed as mean between group differences with 95% confidence intervals. Analyses will be performed with intention-to-treat. Physical activity enjoyment will be measured at 8 time points and will be analyzed using mixed effects modelling. This type of analysis is useful for assessing longitudinal change and can accommodate missing data. Exploratory analyses of all secondary outcomes disaggregated by sex and gender will also be conducted.

### Qualitative Analysis

Data analysis will occur alongside data collection to allow for further refinement of the interview guide as needed. Audio recordings, transcripts, online surveys, and field notes will be stored locally in a secure computer. The responses to the anonymous online surveys will be similarly stored. Qualitative content analysis will be used to analyze the data. This a dynamic form of analysis that aims to summarize the informational content of the data.^44^ The process will involve coding data, recording insights/reflections, identifying similarities and differences, deciding on a some generalizations that hold true for the data, and examining these in light of existing knowledge.^43,65^ Coding, broadly refers to a process of sorting the data into smaller pieces of data to enable further sorting into patterns, relationships, and findings.^66^ This will largely be an inductive process, where codes are data-derived (systematically applied but generated from data itself).^44^ Basic word-processing and spreadsheet software will be used for analysis. Another member of the research team (JR) will review and provide feedback throughout this analysis process.

### Ethics

Participants will be provided with an informed consent form in plain language which details the purpose of the study, what will happen during the study, risks, benefits, confidentiality, and the ability to withdraw at any time. This form will be emailed to participants and reviewed with the participant by AM via videoconferencing. Verbal consent will be documented, and a signed paper copy will be obtained from the participant via mail or picked by AM. This trial has received approval from the Hamilton Integrated Research Ethics Board (Project #13879). There is no external funding agency associated with this project.

## Discussion

This pilot utilizes explicit criteria to determine the feasibility of the process (recruitment), resources (adherence, retention), scientific (safety, potential effects)^42^ aspects of the trial combined with qualitative data to explore the acceptability to it’s participants. This is essential to inform a future fully powered trial and further research. There are several novel aspects of this pilot trial, including the participants, intervention characteristics and outcomes investigated which represent significant areas for researchers’ attention.

To our knowledge, there are no current published trials which examine the effects of exercise on community-dwelling older adults who are experiencing PCML following an injury from a slip, trip, or fall. Individuals with PCML report an absence of difficulty but modifications to mobility activities, indicating greater physical functioning than individuals with overt mobility limitations. However, individuals with PCML are at an increased risk for mobility decline compared to individuals without modifications.^6,7^ The PCML period offers significant opportunity to intervene with exercise. The higher functioning levels of these individuals with PCML may offer an increased propensity and ability to engage in higher intensity exercise.

High-intensity functional strength training (HIFST) is rapidly gaining attention as an efficient means to induce benefits from both aerobic and resistance exercise with a shorter total duration.^12^ A recent systematic review and meta-analyses found this type of training demonstrated improvements in both muscle strength and endurance capacity, but only 4 of 17 trials included older participants and none were home-based.^12^ More research is needed regarding the effects of this HIFST in older adult populations and the feasibility of conducting this type of exercise at home with decreased supervision.

HIFST has not been investigated for older adults in a home-based setting or using virtual methods of delivery. However, a recent large RCT found older adults were able to engage in HIIT over a 5-year period without strict supervision (combination of supervised/unsupervised and various settings based on patient preference) or the occurrence of exercise-related cardiovascular events.^67^ Mañas et al., published a systematic review of unsupervised home-based resistance training which also demonstrated safety and positive effects on muscle strength and power in older adults.^56^ The COVID-19 pandemic drastically increased the availability of online exercise opportunities and has highlighted the need for safe, effective home-based exercise programs. However, barriers for older adults to accessing community exercise programs existed prior to the pandemic (transportation, cost, etc.,) and previous research has demonstrated feasibility and effectiveness for exercise programs delivered via videoconferencing.^68,69^

Finally, the cognitive outcomes, enjoyment, and acceptability of high-intensity exercise for older adults have received minimal attention in the literature. Meta-analyses have demonstrated that combined exercise training (aerobic and resistance) is superior to aerobic exercise alone in improving global cognitive function.^70,71^ Therefore, HIFST may offer a valuable and time-efficient exercise option. The HIFST intervention outlined in this protocol will take individuals approximately 35 minutes to complete (inclusive of warm-up and cool-down). While a typical moderate intensity workout including both aerobic and resistance exercise can easily last 1 hour or longer in duration. Cognitive outcomes of high-intensity exercise for older adults are rarely reported in published interventions,^11,13^ therefore the preliminary effects demonstrated in this trial may help to guide future research directions. The primary feasibility outcome of this pilot is adherence, as an effective exercise program is only useful if individuals are able and willing to adhere to its’ components. The measurement of enjoyment and examination of acceptability using both quantitative and qualitative methods will aid in adding important contextual information to explain the adherence rates observed during the trial and inform future recommendations.

## Conclusion

This paper describes the rationale, methods, and planned analyses of a pilot RCT and follow-up qualitative description study investigating the feasibility, preliminary effects on physical functioning, cognitive functioning, and enjoyment, as well as acceptability of a high-intensity functional strength training intervention for older adults who are post-injury from a slip, trip, or fall. If the results of this trial demonstrate feasibility, preliminary evidence of benefits, and acceptability, HIFST may offer a time-efficient exercise option for older adults that can be completed at home and is highly targeted to function.

## Data Availability

not applicable (protocol)

## Appendix A: Sample Size Calculation

Sample Size calculation

**Proportion of Successes expected related to adherence = 70%**

Margin of error at 0.20

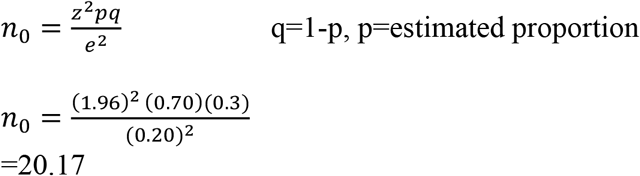

Drop out 15%=20.17*1.15=23

Drop out 20%=20.17*1.2=24

**24 participants** rounded up to an even number to allow for equal numbers between groups

## Appendix B: Outcome Measures

| Outcome Measure | Purpose | Description and Scoring | Interpretation | Examples of Measurement Properties in Older Adult Samples |
| --- | --- | --- | --- | --- |
| 4 m walk test<sup>28</sup> | Assesses physical functioning via usual gait speed | Time (s) required for participant to walk 4 meters at usual speed. Gait speed is calculated in meters/second.<sup>28</sup> | normative values community-dwelling older adults, mean speed in meters/second (sd): men 50-59 yrs. 1.16(0.20), women 1.15(0.20), men 60-69 yrs. 1.16(0.22), women 60-69 yrs. 1.05(0.22), men 70-79 1.07(0.24), women 70-79 yrs. 0.99(0.22), men 80-85 0.97(0.20), women 80-85 0.95(0.24)<sup>28</sup> | test-retest reliability: ICC=0.406<sup>28</sup><br>Videoconferencing compared to face-to-face: ICC=0.77<sup>72</sup> |
| 2 minute step test (TMST)<sup>29</sup> | Assesses physical functioning: exercise capacity | Stepping in place to minimum height (midway between patella and iliac crest) for 2 minutes; score number of times right knee reaches target<sup>29</sup> | high-active older adults mean steps(sd) 95.8(15.7), low active 72.8(18.4), 60-69 yrs. 100.4(9.0), 70-79 yrs. 92.6(16.0), 80-89 yrs. 83.5(22.6)<sup>29</sup> | test-retest reliability: ICC=0.90<sup>29</sup><br>correlation with 1-mile walk time r=0.73<sup>29</sup> |
| dual-task cognitive timed up and go (TUG-COG)<sup>30,31</sup> | Assesses physical functioning (mobility and balance) and dual-task ability | Time (s) required for participant to stand from a chair, walk 3 meters, turn and walk back to sit down (TUG) while counting backward by threes from a randomly selected number between 70-100.<sup>30</sup> | healthy older adults: mean time TUG 8.4 s, mean TUG-COG 9.7-9.8 s,<sup>30,31</sup> cut-offs: identifying fallers 15s<sup>31</sup> | test-retest reliability TUG-COG: ICC=0.98, interrater reliability TUG-COG ICC=0.94<sup>30</sup><br>Videoconferencing compared to face-to-face for TUG: ICC=0.83<sup>72</sup> |
| 30 -second chair stand test<sup>29</sup> | Assesses physical functioning; lower body strength | Number of full stands completed in 30 seconds from chair with arms folded across chest.<sup>29</sup> | High-active older adults mean number of stands (sd) 13.3(2.8), low-active 10.8(3.6), 60-69 yrs. 14.0(2.4), 70-79 yrs. 12.9(3.0), 80-89 yrs. 11.9(3.6)<sup>29</sup> | test-retest reliability ICC=0.89<sup>29</sup><br>correlation with 1RM leg press, r=0.77<sup>29</sup><br>Videoconferencing compared to face-to-face: ICC=0.97<sup>72</sup> |
| Preclinical Mobility Limitation (PCML) Scale based on Mänty<sup>32</sup> | Assesses PCML for 3 tasks (walking 0.5km, walking 2.0 km and climbing one flight of stairs)<sup>32</sup> | PCML is classified no reported mobility difficulty and at least 1 modification in task frequency or performance<sup>32</sup> | PCML increased risk of major mobility disability at 2 year follow-up<sup>32</sup> | unknown |
| California Older Adult Stroop Test (COAST)<sup>33</sup> | Assesses executive functioning; inhibitory control<sup>16</sup> | Involves reading congruent (same colour and word) and incongruent (mismatched colour and word) colour-word pairs. The COAST is designed for older adults and uses fewer items (50) per task, larger font and more easily distinguished colours. Scored on time to completion, uncorrected and corrected errors.<sup>33</sup> | Longer time, greater errors greater limitation in inhibitory control. Time to completion healthy older adults(n=86) mean(sd) color/word interference task 65.56s(21.1) vs. individuals with dementia (n=51) 179.10s(70.6)<sup>33</sup> | test-retest reliability for time to completion (healthy and diabetic older adults) correlations r=0.711-0.907,<sup>33</sup> individuals with dementia r=0.82-0.95<sup>33</sup> |
| Oral Trail Making Test (OTMT)<sup>34,35</sup> | Assesses executive functioning; task shifting (mental flexibility), attention, working memory and processing speed<sup>34,35</sup> | Part A involves counting from 1-25 as quickly as possible, and Part B alternating between counting and the reciting the alphabet (e.g., 1-A, 2-B, etc.) until the examiner says stop (number 13).<sup>34,35</sup> Scored by length of time for each component, errors and can calculate difference between two components<sup>34,35</sup> | Longer time and greater errors = worse performance. Possible impairment cut-off (9<sup>th</sup> percentile), Part A; 49-69yrs=9.6s, 59-79yrs=9.1s, 69-90yrs=9.1s, Part B; 49-69=56s, 59-79=69.7s, 69-90=73s<sup>35</sup> | Oral Part B correlation with Written Part B r=0.62<sup>35</sup> |
| Digit Symbol Substitution Test (DSST)<sup>36,37</sup> | Assesses processing speed, executive functioning; attention, planning, working memory<sup>14,36</sup> | Participants given a list of digits and symbols in which each symbol corresponds to a digit then must write down appropriate symbol on a list of digits as fast as possible.<sup>14</sup> Scored by number of boxes completed correctly in 90 seconds.<sup>37</sup> | Lower scores associated with increased odds of subclinical and clinical disorders of cognition, mobility and mood; mean scores(sd), no cognitive disorder 41.7(12.1), subclinical 28.3(11), clinical 19.2(9.9), no mobility disorder 43.8(12.60), subclinical 37.5(13.5), clinical 28.2(12.8)<sup>37</sup> | test-retest reliability in adults 16-81 yrs. $r=0.89^{73}$ |
| 8-item Physical Activity Enjoyment Scale (PACES)<sup>38</sup> | Assess enjoyment of current physical activity | Self-reported 8-item (7 bipolar point scale) adapted for older adult population<sup>38</sup> from original 18-item measure<sup>74</sup> | Scores 8-56 (higher scores = greater enjoyment)<sup>38</sup> | internal consistency: 8-item developed through confirmatory factor analysis in older adults, strong correlation with original 18-item ($r=0.97-0.98$)<sup>38</sup> |
| Feeling Scale (FS)<sup>39</sup> | Assesses affective response to exercise<sup>39</sup> | Self-reported feelings while participating in exercise on an 11-point scale. Completed during or immediately after exercise.<sup>39</sup> | +5 very good to -5 very bad<sup>39</sup> | unknown |
| Self-Efficacy for Exercise (SEE) Scale<sup>62</sup> | Assesses self-efficacy for exercise<sup>62</sup> | Self-reported confidence for exercise when faced with barriers. 9 item scale (10 point bipolar scale).<sup>62</sup> | Scores 0-90 with higher scores = higher self-efficacy | internal consistency: $\alpha=0.92^{62}$ |
| Activities-specific Balance Confidence (ABC) Scale<sup>63</sup> | Assesses balance confidence<sup>63</sup> | Items scored 0-100% (100% complete confidence), Total score/16(number of items) to reach % of self-confidence | 0% (no confidence) to 100% (complete confidence)<br>Older adults with high mobility mean score 80.9, low mobility: 38.3<sup>63</sup><br>>80%= high level of physical functioning, 50-80%= moderate level, <50%=low level<sup>75</sup> | internal consistency: Cronbach's $\alpha=0.96^{63}$<br>test-retest reliability $r=0.92^{63}$ |

m=meters
m/s=meters/second
s=seconds
ICC=intraclass coefficient
yrs.= years
kg=kilogram
sd= standard deviation
r=correlation coefficient
LoA= limits of agreement

